# Inequality in access to health and care services during lockdown – Findings from the COVID-19 survey in five UK national longitudinal studies

**DOI:** 10.1101/2020.09.12.20191973

**Authors:** Constantin-Cristian Topriceanu, Andrew Wong, James C Moon, Alun D Hughes, David Bann, Nish Chaturvedi, Praveetha Patalay, Gabriella Conti, Gabriella Captur

## Abstract

**Background:** Access to health services and adequate care is influenced by sex, ethnicity, socio-economic position (SEP) and burden of co-morbidities. However, it is unknown whether the COVID-19 pandemic further deepened these already existing health inequalities.

**Methods:** Participants were from five longitudinal age-homogenous British cohorts (born in 2001, 1990, 1970, 1958 and 1946). A web and telephone-based survey provided data on cancelled surgical or medical appointments, and the number of care hours received during the UK COVID-19 national lockdown. Using binary or ordered logistic regression, we evaluated whether these outcomes differed by sex, ethnicity, SEP and having a chronic illness. Adjustment was made for study-design, non-response weights, psychological distress, presence of children or adolescents in the household, keyworker status, and whether participants had received a shielding letter. Meta-analyses were performed across the cohorts and meta-regression evaluated the effect of age as a moderator.

**Findings:** 14891 participants were included. Females (OR 1·40, 95% confidence interval [1·27,1·55]) and those with a chronic illness (OR 1·84 [1·65-2·05]) experienced significantly more cancellations during lockdown (all *p*<0·0001). Ethnic minorities and those with a chronic illness required a higher number of care hours during the lockdown (both OR ≈2·00, all *p*<0·002). Age was not independently associated with either outcome in meta-regression. SEP was not associated with cancellation or care hours.

**Interpretation:** The UK government’s lockdown approach during the COVID-19 pandemic appears to have deepened existing health inequalities, impacting predominantly females, ethnic-minorities and those with chronic illnesses. Public health authorities need to implement urgent policies to ensure equitable access to health and care for all in preparation for a second wave.

## INTRODUCTION

On 11 March 2020, the World Health Organization declared the novel severe acute respiratory syndrome-coronavirus-19 (SARS-CoV-2, also known as COVID-19) outbreak a global pandemic. As the United Kingdom (UK) was facing a surge of new cases, the government-imposed lockdown restrictions across England, Scotland and Wales on 23 March 2020 in order to limit the spread of the virus. Although the restrictions were gradually relaxed, the most widely accepted end-date of the lockdown is considered to be the 4^th^ of July 2020 when non-essential businesses such as bars, restaurants opened. Delivery of routine care across the UK National Health Service was hampered by the pandemic crisis and the lockdown.

Access to health services and adequate care has previously been shown to be influenced by sex, ethnicity, socio-economic position (SEP) and burden of co-morbidities^1,2^. However, it is unknown whether access to health and care services during the COVID-19 pandemic differed by these factors, potentially further widening already existing health inequalities^3^. Evidence from previous pandemics suggests this possibility, but data is missing in the context of COVID-19 currently. To answer these questions, a web-based survey was sent to participants in five UK national longitudinal studies, spanning multiple generations; data were collected during the core UK lockdown, between 2^nd^ of May and 1^st^ of June 2020. We investigated the number of participants having a cancelled surgical or medical appointment and the number of care hours received for self or other household members over a week during the lockdown. We analyzed how these outcomes varied by already established factors contributing to health inequalities. The importance of cancellations stems from the potential consequences of healthcare deprivation, while the number of weekly care hours has been shown to predict admission to long-term care facilities especially in the older population^4^.

## METHODS

### Study design

The five UK national longitudinal studies were: National Study of Health and Development Study (NSHD)^5^, National Child Development Study (NCDS)^6^, 1970 British Cohort Study (BCS70)^7^, Next Steps (NS)^8^ and Millennium Cohort Study (MCS)^9^. NSHD participants were born in 1946, NCDC in 1958, BCS70 in 1970 and MCS in 2000-2002 and all participants were followed-up from birth (all birth cohorts), while NS is a longitudinal cohort study whose participants, born in 1989-1990, have been followed-up from adolescence. The cohorts have been extensively followed up with periodic assessments which have been described elsewhere. During the COVID-19 pandemic (May 2020), an identical online questionnaire, which measured demographic, behavioral and health variables, was sent to each participant from each cohort. The questionnaire was designed to explore the health, care, social, economic, behavioral and psychological consequences of the COVID-19 pandemic. The questionnaire format was multiple choice, but participants were also allowed free text entry to describe their experience in their own words. Ethical approval was obtained from the relevant committees.

### Outcomes

Cancelled surgery, medical procedures or other medical appointments were recoded as a binary variable (yes/1 or no/0). The number of hours of help received for self or other household member in a week during lockdown was recorded in six categories: 0, 1-4, 5-9, 10-19, 20-34 or 35+ hours.

### Exposures

Sex was recoded as 0 = male and 1 = female, while ethnicity was recoded as 0 = non-White and 1 = White. As NSHD, NCDS and BCS70 consist mostly of White participants, ethnicity data was examined only for the NS and MCS cohorts. Highest educational attainment and financial difficulties prior to COVID-19 were used as a proxy for adult SEP. Highest educational attainment was categorized as: degree/higher, advanced-level exam/diploma, ordinary-level exam/general certificate of secondary education or none. Financial difficulties before lockdown were self-rated using the following options: managing comfortably, all right, getting by and difficult. As many MCS participants were still undertaking education and financially dependent on their families, their parents’ highest education and financial difficulties were used. Childhood social class has also been recorded according to the UK Office of Population Censuses and Surveys Registrar General’s social class resulting in six categories: professional, managerial and technical, skilled non-manual, skilled manual, partly-skilled and unskilled.

Participants were asked to report whether they had a long-standing illness (yes/no). In addition, the name of the chronic illness was also recorded. The number of hours of help received for self or another household member in a week before the pandemic was recorded as above. Whether the participant had received a shielding letter was also noted (yes/no). The presence of children aged less than 16 years in the household as well as the self-reported presence of psychological distress during lockdown were recorded (yes/no). The presence of psychological distress was defined as a score of four or over in the General Health Questionnaire (GHQ-12^10^). Keyworker status was self-rated based on whether participants’ work was classified as critical to the COVID-19 response.

### Statistical analysis

Statistical analysis was performed in R (version-3.6.0). Frequency distribution of continuous data were assessed visually using histograms. Categorical variables were expressed as counts and percent for each available category. Within each cohort, childhood SEP, highest educational attainment and financial difficulties were converted into cumulative rank probabilities (ridit scores) to quantify the difference in outcomes comparing the lowest with highest SEP (i.e, the relative indices of inequality)^11^. Models containing all socio-economic variables were assessed for multicollinearity via the variance inflation method. As childhood SEP was multicollinear with the other two, had the least amount of missing data, and as it could impact on adult behaviors and health outcomes independently of adult SEP^12^, it was used in subsequent analysis. However, we additionally report the results from the analyses using SEP based on highest educational attainment, and financial difficulties respectively.

Separate regression models were using sex, ethnicity, SEP and presence of chronic illness as predictors of cancelled appointments or number of care hours needed during lockdown. Generalized linear models with logit link were employed to predict cancelled appointments, while ordinal logistic regression was used to predict the number of care hours needed. The proportional odds assumption for ordinal logistic regression was tested using a Brant test^13^. The analysis was weighted to reduce biases due to missing data. Weights were constructed from logistic regression models predicting the response during the COVID-19 data sweep using demographic, socioeconomic, household and individual predictors of non-response at previous data collection points^14,15^. We also used weights to account for the stratified survey designs of 1946, 1990 and 2000-2002 cohorts^16^. Predictors were included sequentially one at a time. Sex analyses were adjusted for these survey non-response weights and for receipt of a shielding letter. All other analyses were similarly adjusted, but ethnicity analyses were additionally adjusted for sex, SEP analyses additionally adjusted for sex and ethnicity, and chronic illness analyses additionally adjusted for sex, ethnicity and SEP.

Gender differences were further evaluated by adjusting for children less than 16 years in the household and psychological distress during the lockdown. As females are more likely to have a chronic disease, gender differences were also evaluated after adjustment for the presence of a chronic disease^17^. Ethnicity differences were further explored by adjusting for key worker status as ethnic minorities have been reported to be over-represented as key workers in the literature^18^.

Cohort-specific analyses were conducted initially. Meta-analyses were then performed across the cohorts, only if there was a significant result in at least one of the cohorts. Heterogeneity was evaluated using Cochran’ Q test and I^2^ statistic. As smaller samples have more sampling errors in their effect estimate, larger effect size might emerge^19^. Thus, funnel plot asymmetry was evaluated using Egger’s test. Meta-regression was conducted with age/cohort as a moderator in order to determine whether it was a source of heterogeneity. As the associations between age and our outcomes are likely to be non-linear based on visual inspection, we performed the meta-regression using restricted cubic splines modelling^20^.

We ran sensitivity analyses in which we: (1) simulated a complete case analysis through multiple imputation to verify the reliability of observed sex-related differences as the majority of our respondents were female. Using the predictive mean matching method, we have generated 5 complete data sets^21^ and performed a pooled regression. The models were not further adjusted for non-response weights; (2) adjusted the number of care hours during lockdown analyses for the number of care hours before the pandemic; (3) explored possible deviations from the proportional odds assumption via multinomial logistic regression with the number of care hours grouped into Never (0 hours), Low (1-9 hours) and High (10 hours+).

## RESULTS

Overall 15291 participants (45% of the combined cohorts’ participants) responded to the COVID-19 survey as follows: 1241 out of 1842 (NSHD), 5205 out of 8943 (NCDS), 4247 out of 10458 (BCS), 1921 out of 9380 (NS) and 2677 out of 9909 (MCS). Being female, with higher educational attainment, higher income and better self-rated health were associated with higher response rates^16^.

Any participant who lacked data for at least one outcome variable was removed leaving 14891 participants that were included in the final analysis (characteristics summarized in **Table 1**). Overall, included participants were more likely to be female, over 50 years of age and of higher educational attainment. Older participants were more likely to have a chronic illness, receive a shielding letter, experience a cancelled appointment and require more care hours during lockdown. The chronic illnesses recorded spanned a variety of medical systems. Across all cohorts, the most prevalent conditions were high blood pressure (10·9%), recurrent back problems (9·9%), asthma (8·9%) and depression (8.6%)

**Table 1.**
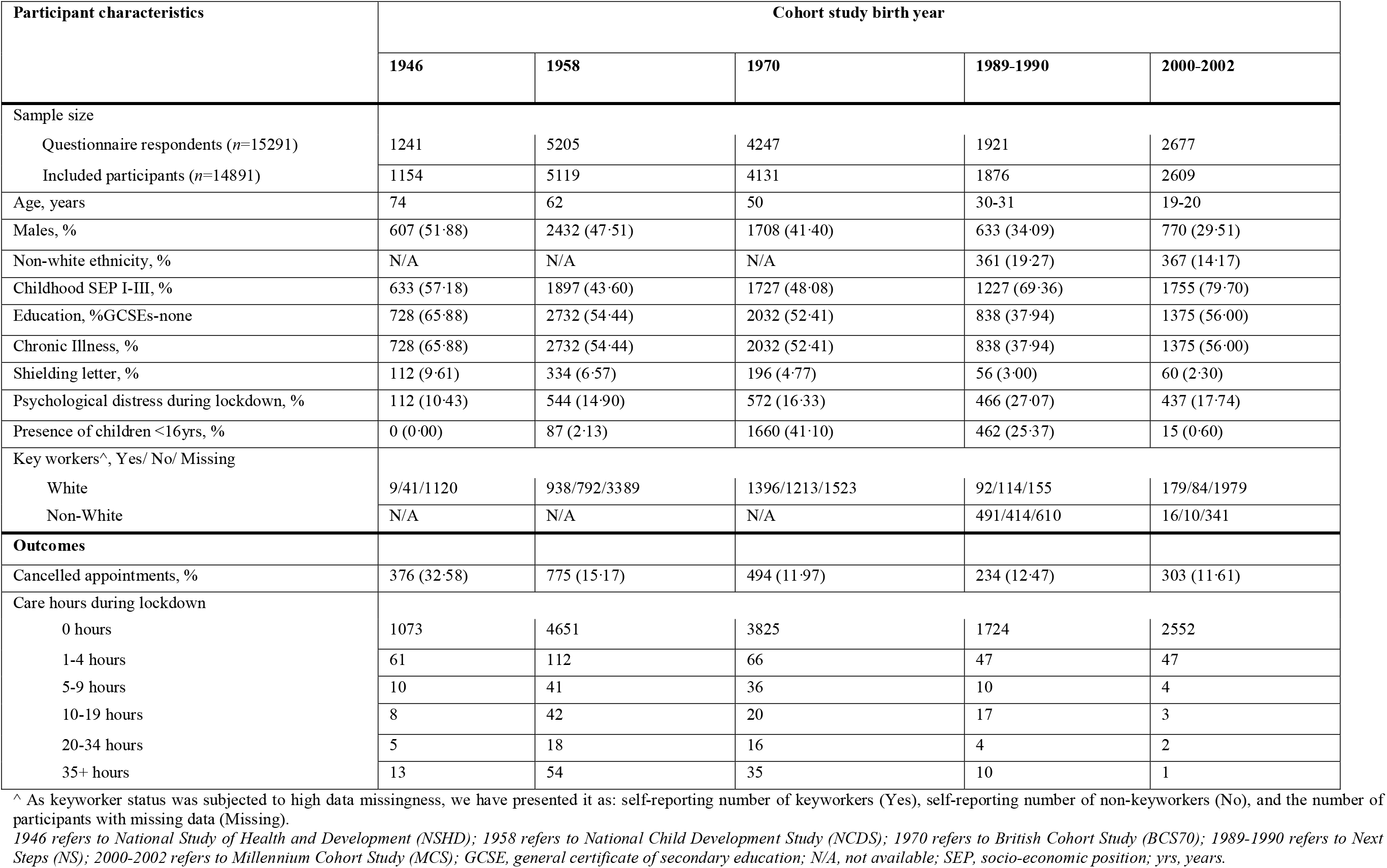
Characteristics of participants by cohort.

### Cancelled surgery, medical procedures or medical appointments during lockdown

In all cohorts except NSHD, female sex was associated with higher odds (ORs range 1·20–2·29, all *p*<0·021) of cancelled surgery, medical procedures or medical appointments (**Table 2**). Adjusting for the presence of children less than 16 years old (**Supplementary Table S1**) and the self-rated presence of psychological distress during lockdown (**Supplementary Table S2**) attenuated the regression coefficients in most cohorts, but sex differences persisted. All the sex differences persisted after adjusting for the presence of a chronic illness, but most coefficients were attenuated (**Supplementary Table S3**). The meta-analysis revealed a pooled OR of 1·40 (95% confidence interval [CI] 1·27, 1·55) in the absence of funnel plot asymmetry (Egger test, *p* = 0·376, **Table 3**). However, there was considerable heterogeneity between the cohorts (I^2^ = 85·78%, *p*<0·0001). In each of the cohorts and in the meta-analysis, presence of a chronic illness at baseline was associated with higher odds (pooled OR 1·84 [1·65, 2·05]) of experiencing a cancelled event. The meta-analysis revealed no heterogeneity (I^2^ = 0·00%, *p* = 0·422) and no evidence of Funnel plot asymmetry when using the standard error as the predictor (Egger test *p* = 0·092). Ethnicity and SEP were not associated with cancellations in any of the cohorts. Age was not significant in the meta-regression (**Supplementary Table S4**). A visual representation of the cancelled surgery, medical procedures or medical appointments by sex, ethnicity and the presence of chronic illness across the 5 UK cohorts is presented in **Figure 1**.

**Table 2.** Association of sex, ethnicity, SEP and the presence of chronic illness with cancelled surgery, medical procedures or other medical appointments during lockdown.

| Cohort study birth year | Sex<sup>^</sup> |  | Ethnicity* |  | Socio-economic position<sup>†</sup> |  | Chronic Illness<sup>§</sup> |  |
| --- | --- | --- | --- | --- | --- | --- | --- | --- |
|  | OR (95% CI) | *p*-value | OR (95% CI) | *p*-value | OR (95% CI) | *p*-value | OR (95% CI) | *p*-value |
| **1946**<br>(*n*=1170) | 0.97<br>(0.76, 1.25) | 0.827 | N/A | N/A | 1.39<br>(0.90, 2.16) | 0.138 | 1.74<br>(1.28, 2.36) | **0.0004** |
| **1958**<br>(*n*=5073) | 1.20<br>(1.03, 1.40) | **0.021** | N/A | N/A | 1.05<br>(0.78, 1.41) | 0.753 | 2.15<br>(1.76, 2.62) | **<0.0001** |
| **1970**<br>(*n*=4099) | 1.83<br>(1.47, 2.26) | **<0.0001** | N/A | N/A | 1.05<br>(0.73, 1.51) | 0.786 | 1.77<br>(1.42, 2.21) | **<0.0001** |
| **1989-1990**<br>(*n*=1849) | 1.70<br>(1.23, 2.35) | **0.001** | 1.25<br>(0.86, 2.37) | 0.255 | 1.45<br>(0.88, 2.41) | 0.154 | 1.59<br>(1.18, 2.13) | **0.002** |
| **2000-2002**<br>(*n*=2605) | 2.29<br>(1.65, 3.19) | **<0.0001** | 1.03<br>(0.73, 2.31) | 0.885 | 1.05<br>(0.66, 1.67) | 0.836 | 1.71<br>(1.30, 2.25) | **0.0001** |

All analyses used generalized linear models with logit link. Significant *p*-values are highlighted in bold.
<sup>^</sup> Sex was coded as 0=male and 1=female; adjustment was made for survey non-response weight and shielding letter.
\* Ethnicity was coded as 0=non-White and 1=White; adjustment was made for survey non-response weight, shielding letter and sex. All participants in NSHD, NCDS and BCS were White, so ethnicity was not examined.
<sup>†</sup> Socio-economic position was coded using childhood social class from 1=managerial to 6=unskilled, but rdit scores were used; adjustment was made for survey non-response weight, shielding letter, sex and ethnicity.
<sup>§</sup> Chronic illness was coded as 0=absent and 1=present; adjustment was made for survey non-response weight, shielding letter, sex, ethnicity and SEP.
OR, odds ratio; CI, confidence interval. Other abbreviations as in **Table 1**.

**Table 3.**
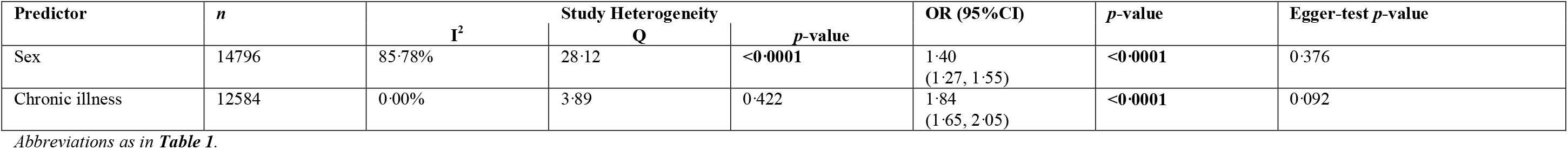
Meta-analysis for the respective association of sex and presence of chronic illness with cancelled surgery, medical procedures or other medical appointments during lockdown.

**Figure 1.**
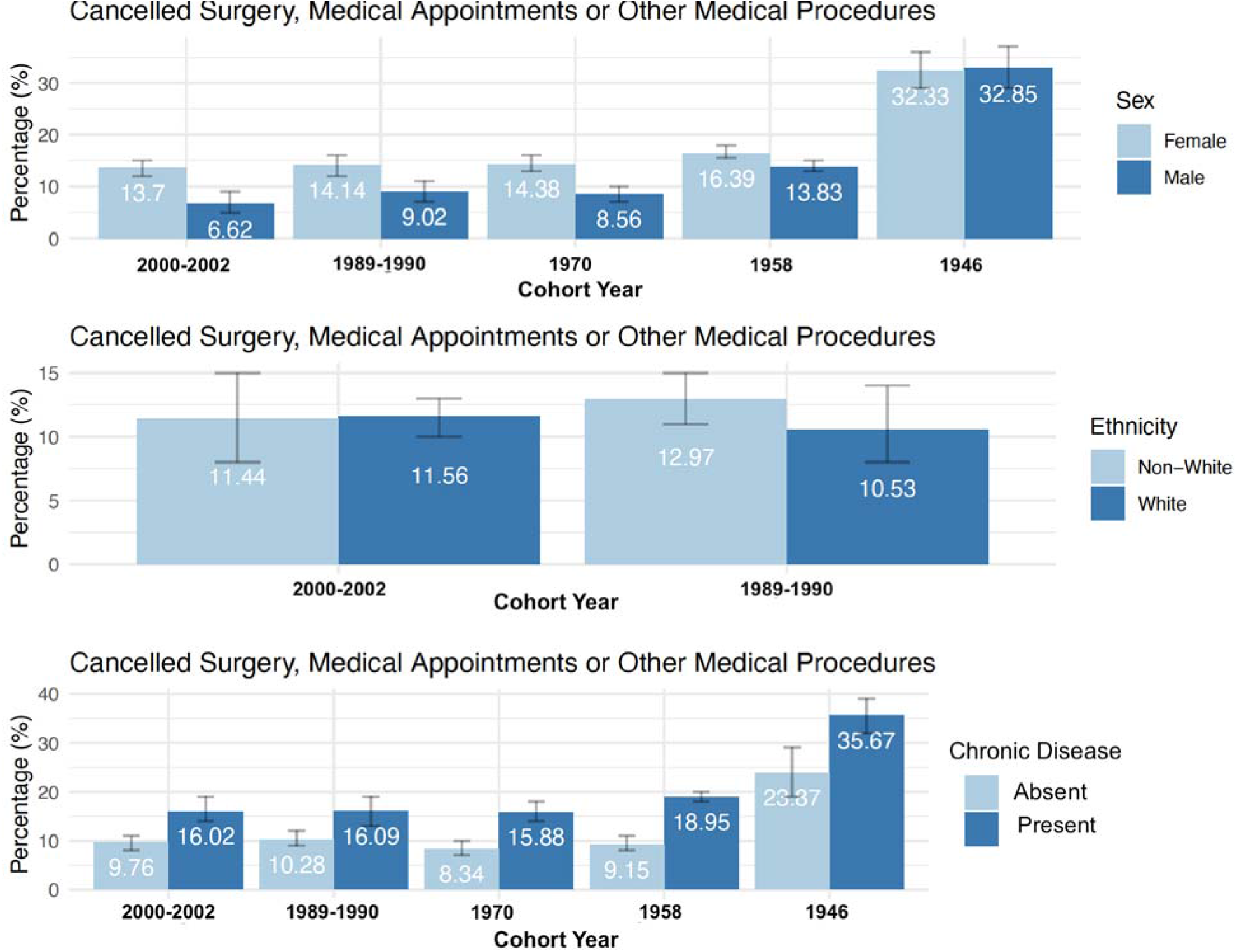
Bar charts illustrating the percentages of cancelled surgery, medical appointments or other medical procedures by sex, ethnicity and the presence of chronic illness across the 5 UK longitudinal cohorts, ordered by increasing age of the cohort from left to right. Error bars representing the 95% confidence intervals are also presented. *1946 refers to National Study of Health and Development (NSHD); 1958 refers to National Child Development Study (NCDS); 1970 refers to British Cohort Study (BCS70); 1989- 1990 refers to Next Steps (NS); 2000-2002 refers to Millennium Cohort Study (MCS).*

### Number of care hours for self or another household member during lockdown

In older cohorts, chronic illness was more prevalent and the association with number of care hours needed was stronger (**Table 4**). In the meta-analysis, higher number of care hours was associated with ethnic minorities (OR 0·53 [0·35, 0·79], I^2^ = 34·17%), and with the presence of chronic illness (OR 2·20 [1·72, 2·56], I^2^ = 13·22%, **Table 5**). After adjusting for keyworker status, significant associations persisted (**Supplementary Table S5**). Sex and SEP were not associated with the number of care hours needed during lockdown. There was no evidence that age contributed to the heterogeneity between cohorts from the meta-regression (**Supplementary Table S1**). Visual representation of the data is provided in **Figure 2**.

**Table 4.** Association of sex, ethnicity, socio-economic position and the presence of chronic illness with number of care hours during lockdown.

| Cohort study<br>birth year | Sex<sup>^</sup> |  |  | Ethnicity<sup>*</sup> |  |  | Socio-economic position<sup>†</sup> |  |  | Chronic Illness<sup>§</sup> |  |  |
| --- | --- | --- | --- | --- | --- | --- | --- | --- | --- | --- | --- | --- |
|  | OR<br>(95% CI) | *p*-value | Brant test<br>*p*-value | OR<br>(95% CI) | *p*-value | Brant test<br>*p*-value | OR<br>(95% CI) | *p*-value | Brant test<br>*p*-value | OR<br>(95% CI) | *p*-value | Brant test<br>*p*-value |
| **1946**<br>(n=1170) | 1.17<br>(0.77, 1.79) | 0.452 | **<0.0001** | N/A | N/A | N/A | 1.17<br>(0.56, 2.45) | 0.683 | 0.329 | 2.20<br>(1.22, 3.99) | **0.009** | 0.192 |
| **1958**<br>(n=4884) | 1.25<br>(0.70, 1.61) | 0.087 | 0.656 | N/A | N/A | N/A | 1.29<br>(0.81, 2.06) | 0.282 | 0.877 | 2.17<br>(1.56, 3.04) | **<0.0001** | 0.860 |
| **1970**<br>(n=3972) | 0.99<br>(0.72, 1.36) | 0.955 | **0.010** | N/A | N/A | N/A | 0.72<br>(0.39, 1.30) | 0.272 | 0.026 | 2.74<br>(1.84, 4.08) | **<0.0001** | 0.244 |
| **1989-1990**<br>(n=1787) | 0.69<br>(0.44, 1.08) | 0.102 | **<0.0001** | 0.44<br>(0.27, 0.72) | **0.001** | **0.005** | 0.87<br>(0.39, 1.94) | 0.727 | **0.0003** | 1.63<br>(1.01, 2.65) | **0.047** | **0.010** |
| **2000-2002**<br>(n=2605) | 0.88<br>(0.49, 1.64) | 0.681 | 0.998 | 0.76<br>(0.38, 1.53) | 0.432 | 0.972 | 2.17<br>(0.77, 6.25) | 0.146 | 0.998 | 1.38<br>(0.75, 2.56) | 0.301 | 0.961 |

All analyses used generalized linear models with ordinal logit link. Socio-economic was recorded using childhood social class. Significant *p*-values are highlighted in bold.
<sup>^</sup> Sex was coded as 0=male and 1=female; adjustment was made for survey non-response weight and shielding letter.
<sup>\*</sup> Ethnicity was coded as 0=non-White and 1=White; adjustment was made for survey non-response weight, shielding letter and sex.
All participants in NSHD, NCDS and BCS were White, so ethnicity was not examined.
<sup>†</sup> Socio-economic position was coded using childhood social class from 1=managerial to 6=unskilled, but ridit scores were used; adjustment was made for survey non-response weight, shielding letter, sex and ethnicity.
<sup>§</sup> Chronic illness was coded as 0=absent and 1=present; adjustment was made for survey non-response weight, shielding letter, sex, ethnicity and SEP.
Abbreviations as in **Table 1**.

**Table 5.** Meta-analysis for the respective association of sex and presence of chronic illness with number of care hours during lockdown.

| Predictor | *n* | *I*<sup>2</sup> | Study Heterogeneity |  | OR (95%CI) | *p*-value | Egger-test *p*-value |
| --- | --- | --- | --- | --- | --- | --- | --- |
|  |  |  | Q | *p*-value |  |  |  |
| Ethnicity | 4371 | 34.17% | 0.218 | 0.218 | 0.53 (0.35, 0.79) | **0.002** | N/A<sup>^</sup> |
| Chronic illness | 12684 | 13.22% | 4.609 | 0.330 | 2.10 (1.72, 2.56) | **<0.0001** | 0.312 |

<sup>^</sup>Egger test was not feasible as only 2 studies recorded ethnicity.
Abbreviations as in **Table 1**.

**Figure 2.**
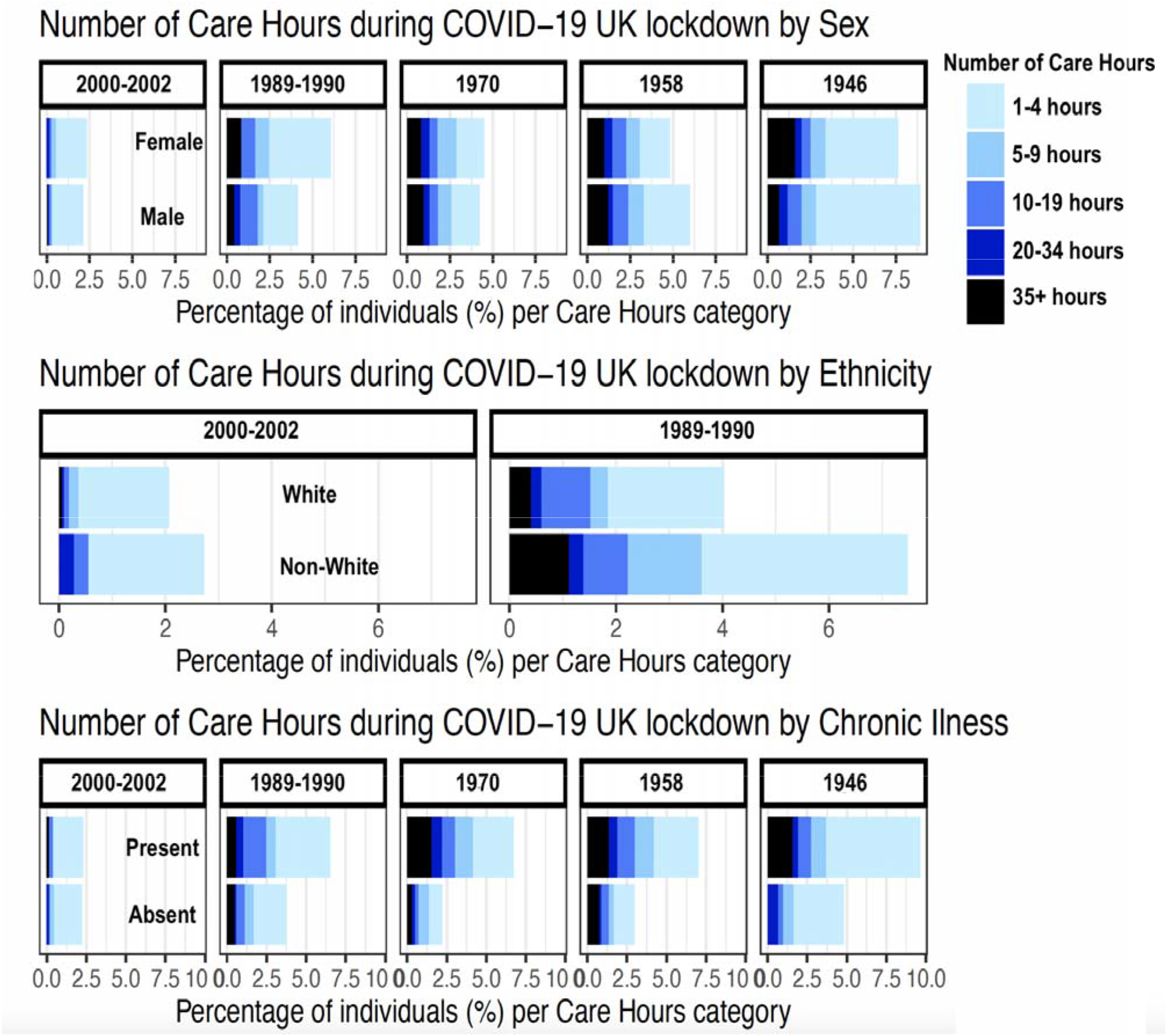
Bar charts illustrating the percentage of participants requiring support based on the number of care hours needed during the UK COVID-19 national lockdown stratified by sex, ethnicity and the presence of chronic illness across the cohorts.

### Sensitivity analysis

Associations between sex and cancelled surgery, medical procedures or other medical appointments persisted after multiple imputation (**Supplementary Table S6**). Adjustment for previous number of care hours attenuated the OR, but the associations mostly persisted (**Supplementary Table S7**). Findings were similar in the multinomial logistic regression when looking at the transition from Never (0 hours) to Low (1-9 hours), but more variability was observed at the transition from Low to High (10 hours +, **Supplementary Table S8**). When using highest educational attainment or financial difficulties before COVID-19 instead of childhood SEP, there were still no significant associations with cancellations (**Supplementary Table S9**) or the number of care hours during lockdown (**Supplementary Table S10**).

## DISCUSSION

These data from five UK national longitudinal studies, at the height of the UK national COVID-19 lockdown in May 2020, indicate that females and those with a chronic illness experienced more cancellations of planned surgical, medical procedural or other medical appointments during lockdown. In addition, ethnic minorities and those with a chronic illness experienced significantly higher care needs during lockdown. Younger generations struggled with access to healthcare and heighted care needs as much as older people. A trend towards increased health inequalities in healthcare access brought about by the COVID-19 pandemic has been observed in United States^22,23^ but to our knowledge this is the first study showing similar effects in the UK.

Chronic illnesses rarely occur in isolation as co-morbidities often co-exist^24^ and interact to produce complex clinical dynamics^25^ (e.g. patients with diabetes are more likely to develop ischaemic heart disease). In addition, patients tend to seek care for all their individual co-morbidities rather than just their major defining condition^26^. Thus, even before the COVID-19 lockdown, this group was vulnerable and required more access to health services, as well as care from family members, friends and care service providers. The pandemic triggered important and unprecedented changes affecting healthcare (which shifted to prioritize COVID-19 patients) and socio-economic dynamics (caused by restricted movement, changes to work patterns and remuneration and unstable housing). Our results show that people with chronic illnesses were twice as likely to have cancelled medical appointments potentially depriving them of vital medical care. They were also twice as likely to require increased number of care hours. Only around 50% of the participants had their care hours expectations met which suggests that a significant proportion were deprived of essential care. These results were observed even after adjustment for demographic factors such as sex, ethnicity, and SEP. Their persistence after adjustment for shielding letter and previous care-hours illustrates their deeply rooted associations with the outcomes. Overall, participants with chronic illnesses received a double-hit with potentially long-lasting effects on their health and wellbeing.

Our study found that females were more likely to experience cancellations in planned surgery, medical procedures or other medical appointments during lockdown. This could be linked to already existing sex inequalities where females adopt a more caring role prioritizing other family members’ needs over their own^27^. Sex inequalities during lockdown could also have widened on account of the added childcare responsibilities including home-schooling, being predominantly undertaken by women. Adjusting for the presence of children under 16 in the household attenuated the regression coefficients suggesting this is a likely contributory factor. Also, in line with sociological gender role models, females would be expected to be more likely to experience anxiety during lockdown. Adjusting for psychological distress also attenuated the regression coefficients in most cohorts. However, one could argue that psychological distress is a collider as it can be independently associated with both sex and cancellations. Lastly, the presence of a chronic illness also contributed to cancellations.

Ethnic minorities were twice as likely to require an increased number of care hours compared to white participants in the younger cohorts. Ethnic minorities are more likely to be in lower paid employment and unstable housing situations, and to rely on public transportation. It is likely that the unstable socio-economic landscape dominated by loss of income, unstable housing, increased psychological distress and reduced community support brought about by the lockdown restrictions adversely impacted these communities. Another explanation could stem from the fact that ethnic minorities are over-represented as key workers^18^. In order to meet the care needs of their communities, they could have been subjected to increased working hours, unusual working environments, stricter work-based controls, and greater exposure to COVID-19, which could have led to both physical and psychological stress. Our data suggests that keyworker status was a contributor as it lowered the observed effects. However, it should be interpreted with caution because of data missingness.

Rather surprisingly, the meta-regression showed that age was not a predictor for cancellations or accentuated care needs, suggesting an age-homogenous effect of the lockdown across the generations. We expected that older generations would be more frail with co-morbidities, and in receipt of a shielding letter when compared to their younger counterparts, and that therefore they would be more likely to need care for activities of daily-living and serial medical appointments to manage their chronic conditions during lockdown. The fact that younger generations (NS and MCS) experienced a substantial number of clinic cancellations and heightened care needs during lockdown is a potentially worrisome indication that the disruption caused by lockdown to education, housing, relationships, employment and finances may have had far reaching effects on the health and wellbeing of young people in the UK.

As pandemics can be characterized by multiple waves, they can last several years^29^. As many experts warn about a second wave in the autumn-winter period, it is vital that public health authorities implement nationally interventions to bolster health and care access. Preliminary data shows that the COVID-19 pandemic further deepened the already existing health inequalities. In addition, the first wave has brought about service disruptions leading to postponed consultations, scans and procedures. As a result, a surge in late-presenting conditions such as cancer is predicted to occur^30^ which will further strain the healthcare system. Thus, public health authorities are faced with the challenges of a finely balanced act: to promote access to healthcare for vulnerable groups on the one hand, whilst minimizing exposure to SARS-CoV-2 infection on the other. Countries without a free healthcare systems where citizens rely on paid insurance such as the United States are in an even more difficult position^31^.

Remote healthcare, also known as telehealth, has been brought forward as a potential solution to the problem of health inequalities in the COVID-19 situation. However, already existing health inequalities predict similar digital inequalities, making telehealth prone to similar problems of unequitable health access provision^32,33^. In order to make telehealth egalitarian, factors contributing to digital inequalities need to be thoroughly understood. These include technical hardware disparities (lack of technological equipment, slower internet connections), digital literacy and access to technical support. Telehealth needs to be modelled to address these digital inequalities from the outset to ensure diffusion of health messages to the most vulnerable groups, and access to timely, effective, safe, culturally-sensitive person-centered healthcare.

Strengths of the study are the implicit age homogeneity of participants enabling age-matching within each cohort as participants were exposed to similar life factors before the national lockdown. Combining five cohorts spanning multiple age groups (19, 30, 50, 62 and 74 years old) enabled a better understanding of how the COVID-19 pandemic has affected different generations. In addition, we have a considerable sample size of more than 14000 respondents and analyses have incorporated non-response weights to account for data missingness which can be problematic in epidemiological studies.

A limitation of the study is data missingness due to low response rates, particularly in younger cohorts, and the small sample size of the older cohorts, particularly in NSHD. However, given the longitudinal nature of the cohorts, all the analyses have been adjusted for via sample weights derived from missingness predictors^14,15^ that would not otherwise be possible for cross-sectional studies. Secondly, we have binarized the ethnicity variable to enable sufficient sample sizes for comparisons which precludes investigation of differences between the diverse ethnic groups which exist in the UK. As the older cohorts (NSHD, NCDS and BCS70) consist of only white participants, we are unable to describe findings for older persons belonging to minority ethnic groups. These individuals may have been most adversely affected by the national lockdown. As self-reported measures were used, the number of care hours needed before and during the lockdown were subject to reporting biases. In addition, single categorical outcome variables do not have the capacity to measure the impact spectrum generated by the cancelled appointments as well as the loss of care hours. Due to study design, the number of care hours were recorded for self or other household member. Moreover, we were unable to separate effects generated by the pandemic from recognized confounders such as seasonal variation in the number of care hours needed, as well as unobserved confounders. The overall prevalence of outcomes differed between cohorts and this can affect the interpretation of ORs, potentially introducing bias between cohort comparisons. Lastly, a limitation of the restricted cubic spline meta-regression is the number of knots per variable as our study included only 5 cohorts^34^.

## CONCLUSION

Individuals with a chronic illness were more likely to experience cancelled healthcare appointments and greater care needs during the UK national lockdown generated by the COVID-19 pandemic. Females experienced reduced access to healthcare, while ethnic minorities required extra care hours. Our results suggest that the pandemic might have widened pre-existing health care inequalities, further depriving already vulnerable and disadvantaged groups of the health and care services which they need. Public health measures should be rapidly implemented to better protect and meet the health and care demands of such at risk groups ahead of a COVID-19 second wave.

## Data Availability

NSHD data is available from: https://www.nshd.mrc.ac.uk/data. Data from the remaining cohorts is available from the UK Data Archive: https://www.data-archive.ac.uk.

## ACKNOWLEDGEMENTS

We are grateful to all the members of our studies for their contribution to this COVID-19 survey and for their ongoing participation in our studies. We thank the Survey, Data, and Administrative teams at the Center for Longitudinal Studies and Unit for Lifelong Health and Ageing, UCL, for enabling the rapid COVID-19 data collection to take place.

## AUTHOR CONTRIBUTIONS

All authors contributed significantly to the design, implementation, analysis, interpretation and manuscript writing.

